# Health literacy and institutional delivery among young Angolan women: a cross-sectional study in three provinces

**DOI:** 10.1101/2025.03.11.25323791

**Authors:** Francisca Van Dunem dos Reis, Arciolanda Macama, Signe Schjøtt, Lucie Laflamme, Barbora Kessel, Gunilla Elise Priebe

**Affiliations:** Faculty of Economics and Management, The Catholic University of Angola, Av. Pedro de Castro Van-Dúnem Loy 24, Luanda, Angola; National School of Public Health, Nova University of Lisbon, Avenida Padre Cruz, 1600-560 Lisbon, Portugal; School of Public Health and Community Medicine, University of Gothenburg, Box 469, 405 30 Gothenburg, Sweden; Department for Global Public Health, Karolinska Institutet, 171 77 Stockholm, Sweden; Institute for Social and Health Sciences, University of South Africa, Pretoria, South Africa

## Abstract

A reduction in maternal mortality remains critical, particularly in sub-Saharan Africa, were significant disparities in access to healthcare affect outcomes. In Angola, almost half of women give birth outside the health system, highlighting the need to increase access to maternity services and information related to women’s sexual and reproductive health and rights (SRHR). The study comprehensively examines young Angolan women’s health literacy in SRHR and its role for institutional delivery. The data were derived from a cross-sectional survey involving 1139 women aged 18–24 years who had given birth, recruited from the provinces of Luanda, Huambo and Lunda Sul. The results included a univariate description of the participants’ socio-demographics and SRHR characteristics, and the barriers and facilitators to institutional delivery they identified. In addition, logistic regression models assessed associations between the participants health literacy levels and the odds for institutional delivery. Findings revealed that more than half of the women delivered in healthcare institutions. These women were comparably economically advantaged, often literate and resided in urban areas. Major barriers identified included lack of transportation and financial resources; facilitators also included companionship and trust in respectful treatment. Regardless of the delivery setting, many women lacked information on fundamental SRHR topics. There were significant associations between health literacy and the site of delivery in all areas considered, except in Lunda Sul. Women who had acquired SRHR information from healthcare professionals, had access to several sources of SRHR information and were acquainted with multiple SRHR topics exhibited higher odds of having had an institutional delivery. The study underscores the need to bolster young women’s access to essential SRHR information and knowledge. However, addressing socio-economic barriers and healthcare limitations concurrently is essential. Multisectoral approaches are vital to ensure widespread access to quality maternity services, thereby ensuring women can safely give birth in quality healthcare settings.

## Introduction

### Women’s right to accessible and appropriate maternity services

Reduction in maternal mortality is an important element of global health and development [1,2]. This particularly true for sub-Saharan Africa, which accounts for 70% of all maternal deaths [3]. The death of a mother has profound consequences, including not only the tragedy suffered by the affected individual, but also the wider family unit, particularly the surviving children [4–6]. A significant proportion of maternal deaths could be averted through utilization of maternal healthcare institutions appropriately equipped for the management of common complications such as haemorrhage, infections and hypertension [2–6]. Empirical evidence indicates that maternal mortality is lower among women who give birth in substandard healthcare institutions compared with women who give birth in non-healthcare settings [7,8].

Thus, the extent to which states fulfil their obligations to make quality maternity services available is a key determinant of the risk of maternal mortality [4–9].

Human rights-based work has identified key factors related to the accessibility and acceptability of healthcare, such as how it is delivered, i.e. whether interactions between care recipients and health professionals are constructive, and whether the health system is designed to accommodate women’s different circumstances [10,11]. Research shows that much work remains to be done in this respect because a wide range of socio-economic factors, including age, level of education, financial resources and transportation resources, are associated with access to maternity services [6,12–16]. The healthcare environment is sometimes not perceived as acceptable. For example, suboptimal quality of antenatal care as well as fear of disrespectful treatment have been identified as barriers for women to giving birth in a healthcare institution [17–19].

### The health promoting potential of health literacy

Another factor influencing the availability and utilization of maternity services is women’s access to information regarding the functionality, timing and rationale for its deployment [13,20,21]. This has been a particular focus of health literacy research; basic, interactive and critical health literacy have been identified as three distinct levels [22]. The former includes access to fundamental information and understanding in a particular area of health, and the latter two include health-related interaction with others and a progressively advanced ability to use health knowledge to take effective action to promote one’s own health [22–27]. Thus, health literacy is understood as a continuum based on the amount of exposure to information, individual capacity, and accessible and acceptable focal points for applying knowledge [22–27].

In addition, health literacy has been described as a stimulant of salutogenic processes, insofar as it can engender self-esteem and a sense of meaningfulness in interplay with other fundamental needs and rights [28]. A fundamental supposition is that advancement towards higher levels engenders heightened autonomy and empowerment; consequently, the magnitude of health literacy exerts an influence on its prospective health ramifications [22–28]. However, even basic health literacy is a prerequisite for understanding oneself and asserting one’s needs in interactions with others, and, accordingly, it has been defined as a fundamental human right and identified as key to positive health development [29,30].

### Health literacy in sexual and reproductive health and rights

A direct causal relationship between health literacy and sexual and reproductive health and rights (SRHR) outcomes has yet to be established conclusively. Nevertheless, several systematic reviews have demonstrated clear associations between health literacy and a range of SRHR-promoting behaviours, including antenatal care-seeking, improved communication with maternal health professionals, and the adoption of practices such as the use of vitamin A supplements and breastfeeding [31–35]. Unfortunately, most of this research has been conducted in countries where socio-economic conditions and health systems are not necessarily linked to increased maternal mortality and limited access to maternity services [36–39]. This presents a particular problem of generalizability, because, as several studies note, the meaning of health literacy emerges in the interactions between the healthcare users, their general life situation and the nature of the health system [23,33,38–41]. It has been recognized that the health system in which individuals of different socio-economic statuses apply their health competencies “has to be health literate, too” [37], i.e. that the successful realization of health literacy-based action presupposes a responsive health system.

The few health literacy studies from regions with under-resourced maternity services report findings similar to those of mainstream health literacy research, namely the potential for a link between health literacy and health promotive actions. For instance, a study conducted in Nigeria demonstrated that adolescents who had a high level of maternal health literacy (defined as knowledge related to pregnancy) were more likely to attend antenatal care visits and adhere to the antenatal tetanus vaccination schedule as prescribed [42]. Similarly, a study from Ghana established a correlation between women’s awareness of pregnancy complications and utilization of maternity services [43]. In addition, research in Botswana has demonstrated that the young people’s source of SRHR-related information is a significant factor. The study found that adolescents who received family planning information from health professionals were eight times more likely to report correct knowledge of condom use compared with those who received information from teachers [44].

This is promising and merits attention, because health literacy, in addition to the advantages mentioned above, has been identified as a component that is relatively easy to influence for the reduction of health inequalities and disparities [31,45–48]. In parallel with this, however, the need to acknowledge not only the potential of health literacy but also its constraints in rectifying entrenched inequalities that emanate from vastly imbalanced opportunities and resource allocation, merits consideration [22,25,30,33]. This necessitates a commitment to enhancing general living conditions, as well as strengthening health systems to ensure the universal realization of SRHR for all women. However, research suggests that the promotion of health literacy, in conjunction with long-term efforts, may serve as an important strategy to the empowerment of women [5,39,42,49–51], thereby facilitating their effective access to healthcare.

### The sexual and reproductive health and rights of women in Angola

The potential for positive development in Angola is closely linked to the situation of the country’s young women; they are a significant segment of its population and are in the nascent stages of societal participation and integration. It is particularly important to invest in young women because over time, stable health behaviours often develop at a young age and SRHR-related actions of a young woman have an impact on her current and future health and that of her children [6].

The importance of addressing these issues is further compounded by the fact that Angola ranks high on gender inequality [52], and Angolan women face even greater barriers than men in several areas related to systemic health inequities, including less access to education, employment and basic healthcare [52–55]. About half of all women still give birth outside the health system and without a health professional present [56,57], a rate that is slightly lower than the average for Sub-Saharan Africa (64%) and among the lowest worldwide [56]. Correspondingly, the country has one of the highest maternal mortality rates in the world, with 1 in 70 women at risk of dying during pregnancy or childbirth [4]. This is attributed in part to a lack of trained medical staff and limited access to maternity services, especially in rural areas [17]. For instance, in Angola, there are around 0.8 hospital beds per 1000 people compared with a global average of 2.9, and only 4% of women are covered by health insurance [52]. Teenage girls are described as being especially disadvantaged in terms of access to healthcare, particularly during pregnancy [55].

Gender-based violence is still a significant concern; more than a third of Angolan women report experiences of physical violence [56–58]. Moreover, fertility rates remain high with an average of 6.2 children per woman, in parallel with limited access to reproductive health education and family planning services [56,57]. Pregnancy between the ages of 10 and 14 years, one of the main risk factors for adverse pregnancy outcomes [6,59,60], is higher in Angola than in any other country [4], making it all the more important to focus on young women in this context. Although official statistics suggest some progress [4,61], independent research shows substantial underreporting of maternal mortality as well as wide socio-economic and regional disparities in access to maternal healthcare and safe family planning methods [62–66].

### Objectives

The study examines the general socio-economic and SRHR situation of young Angolan women in three different provinces with varying levels of institutional development and urbanization. Their perceived barriers and facilitators for institutional delivery are described, and their different levels of health literacy in the SRHR domain, as well as the links between health literacy and institutional delivery, are investigated.

The objective of this study is threefold: firstly, to contribute to knowledge about the living conditions of women at high risk of maternal complications in an under-researched context; and secondly, to enhance the understanding of the potential of health literacy to promote young women’s health. The latter is explored in one of today’s most pressing women’s rights issues, namely the ability to give birth in a healthcare institution if desired.

The importance of childbirth occurring in a healthcare institution with skilled personnel is such that it is considered a vital indicator of the functioning of a health system and, by extension, of a nation’s development [5,9]. The third objective is therefore to highlight the perspectives of young Angolan women, with a view to increasing their incorporation into the development of maternity services. This rests on the premise that the integration of women’s experiences and perceptions into matters pivotal to their lives and societal development is a fundamental human right and is a crucial element in optimizing development strategies.

## Material and methods

This study used a subset of data originating from a larger research project named SADIMA (an abbreviation of the Portuguese Saúde e Direitos das Mulheres em Angola). The subset consists of those women who had ever given birth to a child and reported the place of delivery (S1 Fig).

### Ethics approval and considerations

Ethical approval was obtained from the Ethics Committee Institutional Review Board of the Angolan Ministry of Health (24/C.E.). U/2021, 2021-07-07), the Ethics Committee at the Universidade Católica de Angola (Approvação 153, CEIH 230, 2021-06-30) and the Swedish Ethics Review Authority (Dnr 2022-06393-01, 2023-01-25).

Verbal informed consent was obtained using the DHS model [67]. Each question included an option to decline to answer, ensuring continuous voluntary participation. Ethical considerations characterized all steps of the research process, including the development of the questionnaire, the implementation of data collection, analysis and interpretation of data.

### Setting

Angola is located on the south-west coast of Africa. Despite rich natural resources, as much as half of the population lives in poverty [68]. Angola is one of the ten most economically unequal societies in the world, a variation that is reflected in the three provinces Luanda, Huambo and Lunda Sul [57]. For example, DHS data show that more than two-thirds of women in Luanda are literate, while this was the case for around half of the women in Huambo and Lunda Sul [57]. Similarly, the data showed considerable variations in access to reproductive health services: approximately one-third of women in Luanda and Huambo reported having postpartum consultations within 2 days, whereas fewer than one twentieth of women in Lunda Sul had this experience [57]. Huambo and Lunda Sul are both predominantly rural provinces, and Lunda Sul is the most economically disadvantaged of the three. The capital province of Luanda is highly urbanized [57,69,70].

### Study design

This was a cross-sectional survey-based study.

### Study size

An estimated sample size of 754 young women, calculated based on the latest Angolan Demographic Health Survey (DHS) report at the time [57] of women aged 16–19 years, was further adjusted for an expected response rate of 80% and a design effect of 2, resulting in a target of recruiting at least 1885 young women. This number was distributed across Luanda, Huambo and Lunda Sul proportional to the cube root of the population size of the selected communes (administrative units within municipalities).

### Study participants, recruitment and data collection

The SADIMA project was designed to include all women aged 18–24 years who were living in one of the selected areas at the time of data collection [71]. No up-to-date household data were available; therefore, sampling was done in three steps to ensure variation in terms of socio-economic status and urbanity. First, six communes per province were selected based on an index constructed using principal component analysis (PCA) of five socio-economic indicators relevant to both rural and urban areas [71,72] (two communes with the lowest, middle and highest PCA values were selected in each province). Second, data collection sites were randomly selected in the urban communes using a grid [73]; for the rural communes, the main village and one other randomly selected village were included. Third, participating women were identified using the spin-the-bottle principle [74], i.e. by interviewers going from house to house and offering participation in the study to any eligible woman in the household.

The data collection took place between 1 February and 19 May 2022 and was conducted by trained young, university-educated women. In Huambo and Lunda Sul, the research team collaborated with local Civil Society Organisations (CSOs) to establish regulatory acceptance of the study and identify suitable data collectors (S2 Table). Study information (oral and written) was shared at each site 1–3 days before the data collection to reach eligible women. Female interviewers were used to create a trusting atmosphere. They conducted four interviews per day on average until the specified number for that area was reached. Snacks and drinks were provided during the interviews to prevent participant fatigue, and rural participants were compensated with a package of salt and rice for lost work time.

At the end of each day’s activities, completed surveys were transferred to a Microsoft Excel worksheet without any individual identifiers and stored on a secure hard drive. Controls were conducted daily to ascertain internal consistency, and assessments were carried out at the end of the work week to identify potential interviewer bias.

The number of eligible women who were not included in the study could not be determined because reliable household data were not available. However, information about the eligibility of the household and whether the household participated in the study was recorded. This showed a participation rate of 95% of eligible individuals contacted [71].

### Data collection instrument

The SADIMA project investigates the social determinants of young Angolan women’s SRHR and psychological well-being [71]. The questionnaire was developed between July and December 2021 based on literature searches and key informant interviews with public health experts and CSOs in the three provinces. It was tested for appropriateness and understandability in focus group discussions with female students and data collectors and in interviews with 66 members of the target population. The development process continued until the questions were perceived as easy to understand, respectful, precise and in logical order. Common idiomatic expressions were added in parentheses to improve comprehensibility, and keywords in local languages were recorded on a separate sheet. The final version of the questionnaire included 150 questions in five sections: socio-demographic characteristics, psychological well-being, gender norms, SRHR (pregnancy, menstrual health, family planning and domestic violence), and social capital. It was coded using Adobe In-Design for use on tablets or printed on paper [71].

### Variables

Forty-three questions from the SADIMA questionnaire were included in the current study. These were used to construct a range of variables within four thematic areas: socio-demographic and SRHR characteristics (*n*=16), accessibility and acceptability barriers and facilitators to institutional delivery (*n*=9), health literacy (*n*=7), and institutional delivery (*n*=1) (detailed information on the wording and origin of the variables is given in S3 Table).

The outcome variable, institutional delivery (yes/no), reflects the organization of the Angolan health system. The response options hospital delivery and health centre delivery were combined into the category institutional delivery and the remaining options (health post delivery, home environment delivery, other) were combined into the category non-institutional delivery.

The variables within the socio-demographic and SRHR characteristics theme included province (Luanda/Huambo/Lunda Sul), area of residence (urban/rural), household wealth (upper/lower half), household night hunger (yes/no), formal education (high/medium/low), literate (yes/no), currently in a relationship (yes/no), family planning use (yes/no), aged >18 years at first childbirth (yes/no), given birth to more than one child (yes/no), preference for institutional delivery (yes/no), attended antenatal healthcare check-ups during last pregnancy (yes/no), attended by a healthcare professional at last childbirth (yes/no), complications at last pregnancy or childbirth (yes/no), infant or child mortality (yes/no) and intimate partner violence (yes/no).

Variables were also included to reflect the participants’ views on the barriers and facilitators to institutional delivery, historically and for future scenarios, mirroring the human rights-based AAAQ model’s definitions of healthcare accessibility and acceptability [10,11]. Women who had not given birth in a healthcare institution at their last pregnancy were asked whether or not this was due to accessibility barriers: transportation (yes/no), financial resources (yes/no), companionship (yes/no), knowledge of maternity services (yes/no); and/or acceptability barriers: self-motivation (yes/no), partner or family motivation (yes/no), trust in respectful treatment (yes/no), trust in maternal care quality (yes/no); and other barriers (yes/no). All participating women were also asked what top two factors would facilitate institutional delivery in a potential future pregnancy with the same response options as given for the barriers to institutional delivery.

Finally, the health literacy variables were constructed based on survey questions developed specifically for the SADIMA study because the existing instruments either were not sufficiently contextually relevant [27,38,39,75] or did not cover the comprehensive field of SRHR [32,75–79]. Salient SRHR topics were identified during key informant interviews, via a thorough review of the research and policy literature in the fields of health literacy [22–29,31–35,37,40,41,46,75–79] and SRHR [1,2,5,7,8,10–12,15,19,20,34,43,55,59,62–66,80–83], and by examining the DHS questionnaire for Angola [67] (S3 Table).

Five fundamental and pertinent SRHR topics were identified: fertile period and cycle, pregnancy or childbirth complications, family planning, menstrual health and domestic violence. The first reflected actual knowledge, whereas the subsequent four topics encompassed information acquired about the SRHR topic and how to access products or support if needed. Moreover, the variable for fertility period was ‘acquired knowledge of fertile period and cycle’ (yes/no), whereas the variables for the other topics also captured potential interaction with multiple information sources. Response options for acquired information on pregnancy or childbirth complications; family planning; menstrual health; and domestic violence were Yes, from >1 source/Yes, from 1 source/No. The variable ‘acquired SRHR information from health professionals’ (yes/no) specifically reflected potential interactions with healthcare representatives. The variable ‘acquired SRHR awareness’ (high: 4–5 topic areas/medium: 2–3 topic areas/low: 0–1 topic) included all five topics, indicating the extent of thematic coverage regarding respondents’ SRHR information interactions. These constructs aimed to reflect the breadth of the SRHR field as well as the emphasis on health literacy levels in the literature as being important for their association with health outcomes.

### Statistical methods

The general and SRHR characteristics of the participants were summarized as the number and proportion, according to where the respondents had given birth to their last child. Participants’ responses to questions about barriers and facilitators to institutional delivery and health literacy were summarized per province and type of residential area.

Subsequently, the associations between the health literacy variables and institutional delivery were modelled using logistic regression. These are presented as odds ratios with 95% confidence intervals. Crude and adjusted odds ratios for institutional delivery were calculated for each health literacy variable. The analysis was stratified by province and type of residential area. Models per province included adjustments for literacy, household night hunger, household wealth, type of residential area, age >18 years at first childbirth, and giving birth to >1 child. Models per area of residence included adjustments for the same set of variables except the type of residential area was replaced with province. The statistical significance of associations was assessed using the likelihood ratio test without adjustment for multiple testing. A 5% significance level was used. The analyses were performed using SPSS (version 29.0.0.0) and STATA (17.0).

## Results

For the SADIMA study, 2109 women met the inclusion criteria and consented to participate in the study. Five were excluded because information on age was missing. The current analyses focus on those 1139 respondents (54.1%) who had ever given birth to a child and reported the place of delivery (S1 Fig).

### Study population characteristics

Table 1 presents the socio-demographic and SRHR characteristics of the respondents according to whether or not they had their last childbirth in a healthcare institution. The two groups differed in several respects; higher proportions of women who had delivered in healthcare institutions were from urban areas (59.4% versus 27.8% of those who had given birth in a non-healthcare setting) and exhibited more favourable socio-economic circumstances, including being in the economically advantaged half (61.9% versus 29.7%) and literate (61.2% versus 37.3%). Similarly, for SRHR characteristics, practising any form of family planning and being >18 years of age at first delivery were more common among women who had delivered in a healthcare institution (56.3% versus 37.1% and 57.9% versus 48.7%, respectively).

**Table 1.**
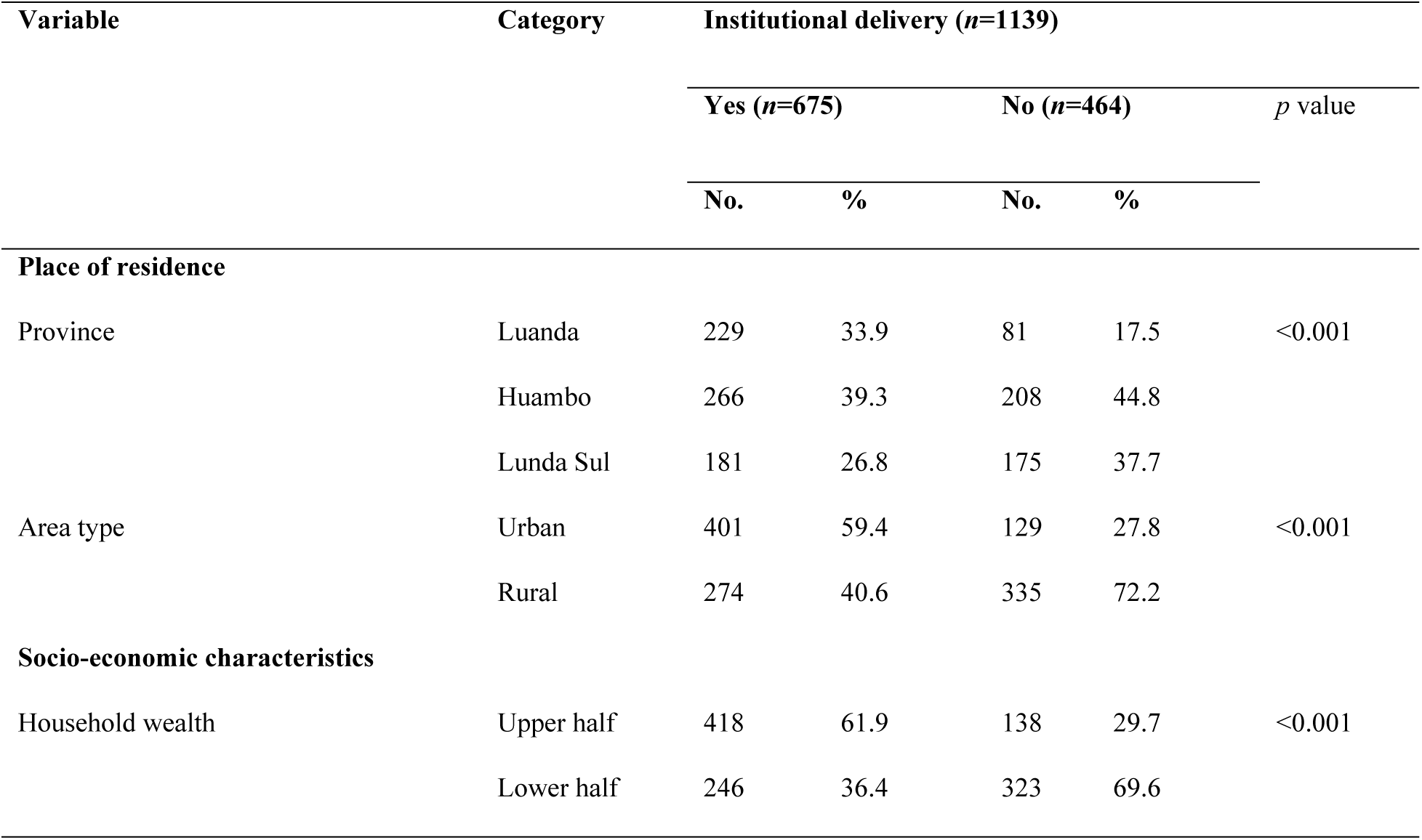
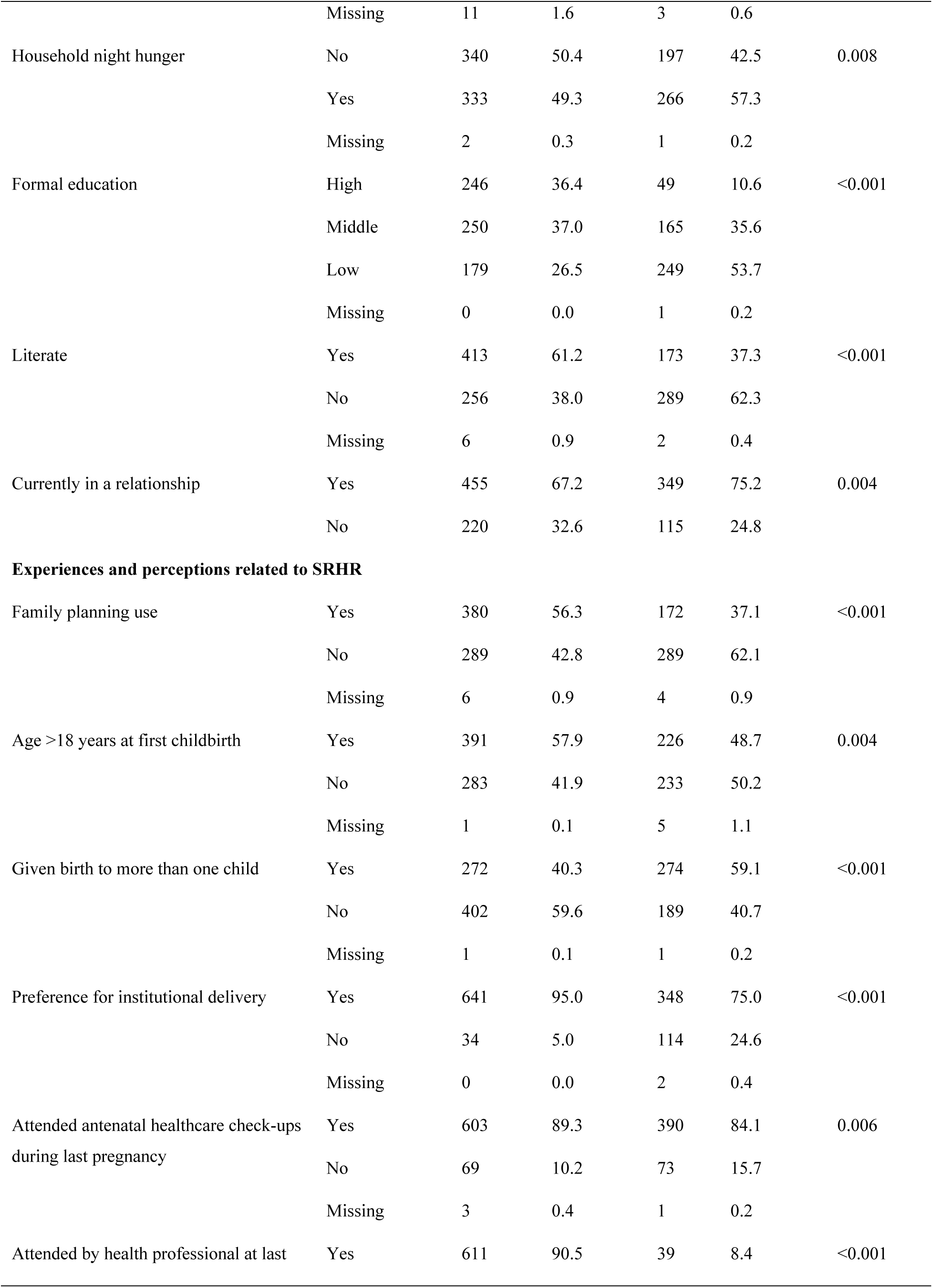
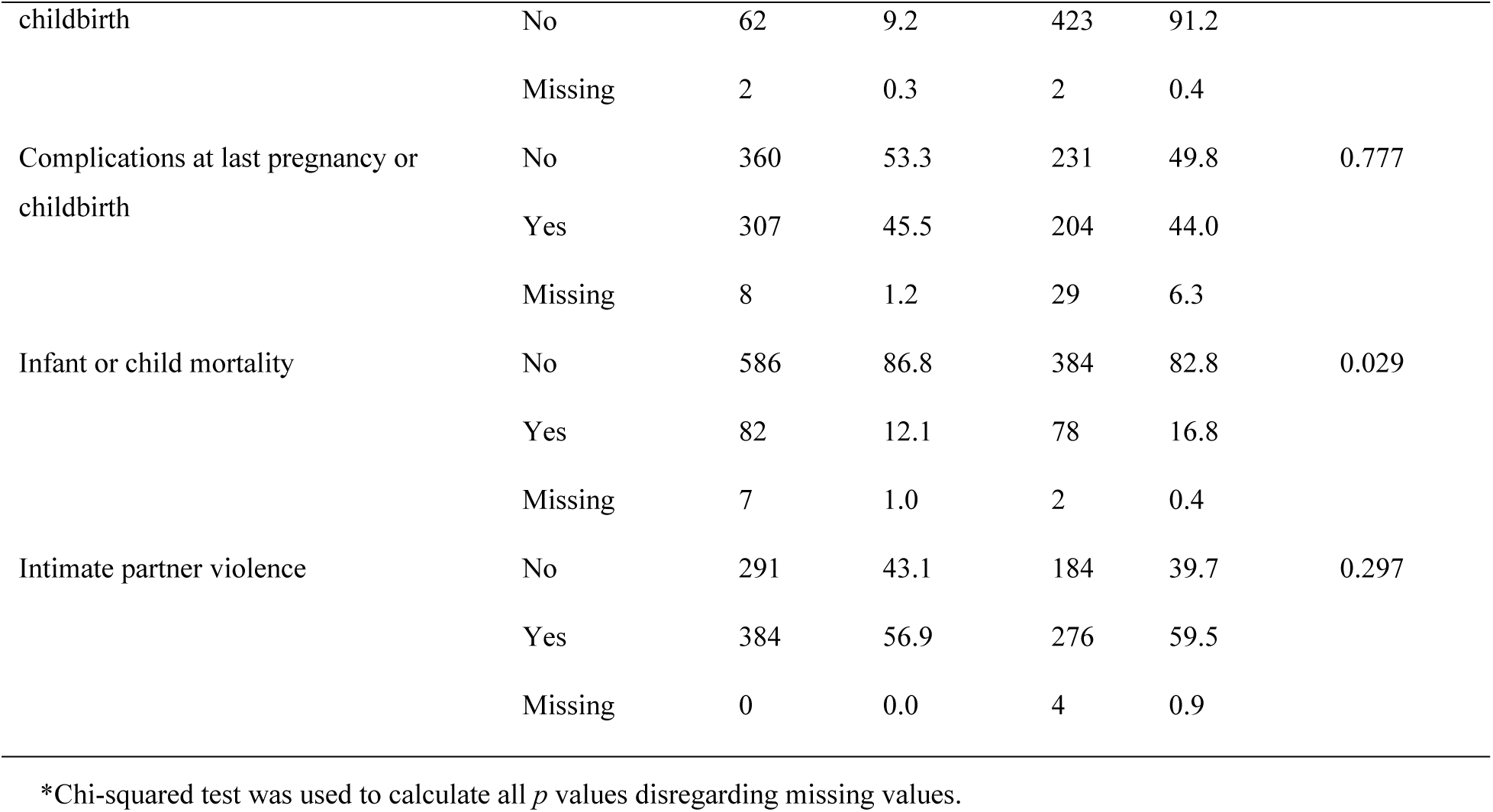
Socio-demographic and SRHR characteristics of participants according to the delivery site.

Most women in both groups indicated that a healthcare institution was their preferred setting for childbirth, although the proportion was higher among those who had given birth in a healthcare institution (95% versus 75%). Other similarities between the groups were a high proportion of women reporting having visited a healthcare institution for antenatal examinations (89.3% and 84.1%), more than 40% having experienced complications during their most recent pregnancy or childbirth (45.5% and 44%), and ∼60% reporting intimate partner violence (56.9% and 59.5%).

### Factors perceived as barriers to institutional delivery at the last childbirth

The women who had not given birth to their last child in a healthcare institution (*n*=464) were asked why this was the case; the responses are presented in Table 2 according to accessibility and acceptability. Lack of suitable transportation was the most frequent barrier mentioned in all settings considered, reaching 40.5% of all women, and was more frequent in Lunda Sul (54.9%) and rural areas (43.9%). Further, the lack of financial resources was the second most frequent accessibility barrier (21.8% of all respondents) and, again, was more prevalent among young women from Lunda Sul (27.4%) and rural areas (25.1%). Regarding access to information, just a few women cited lack of knowledge about where to give birth in a healthcare institution as a barrier (3.7% across all groups of women). Except in Luanda, the acceptability factor most frequently mentioned was self-motivation to seek care (39.4% in Huambo and 33.4% in rural areas).

**Table 2.** Barriers to institutional delivery at last childbirth by province and area of residence.

| Factor | Province of residence |  |  |  |  |  | Area of residence |  |  |  |  |  | Total (n=464) |  |
| --- | --- | --- | --- | --- | --- | --- | --- | --- | --- | --- | --- | --- | --- | --- |
|  | Luanda (n=81) |  | Huambo (n=208) |  | Lunda Sul (n=175) |  | p value | Urban (n=129) |  | Rural (n=335) |  | p value | No. | % |
|  | No. | % | No. | % | No. | % |  | No. | % | No. | % |  |  |  |
| Accessibility barrier |  |  |  |  |  |  |  |  |  |  |  |  |  |  |
| Transportation | 27 | 33.3 | 65 | 31.3 | 96 | 54.9 | <0.001 | 41 | 31.8 | 147 | 43.9 | 0.017 | 188 | 40.5 |
| Financial resources | 12 | 14.8 | 41 | 19.7 | 48 | 27.4 | 0.50 | 17 | 13.2 | 84 | 25.1 | 0.005 | 101 | 21.8 |
| Companionship | 8 | 9.9 | 24 | 11.5 | 26 | 14.9 | 0.458 | 16 | 12.4 | 42 | 12.5 | 0.944 | 58 | 12.5 |
| Knowledge of maternity services | 3 | 3.7 | 10 | 4.8 | 4 | 2.3 | 0.419 | 4 | 3.1 | 13 | 3.9 | 0.705 | 17 | 3.7 |
| Acceptability barrier |  |  |  |  |  |  |  |  |  |  |  |  |  |  |
| Self-motivation | 12 | 14.8 | 82 | 39.4 | 49 | 28.0 | <0.001 | 31 | 24.0 | 112 | 33.4 | 0.047 | 143 | 30.8 |
| Partner/family motivation | 5 | 6.2 | 14 | 6.7 | 21 | 12.0 | 0.111 | 8 | 6.2 | 32 | 9.6 | 0.239 | 40 | 8.6 |
| Trust in a respectful treatment | 9 | 11.1 | 14 | 6.7 | 8 | 4.6 | 0.138 | 10 | 7.8 | 21 | 6.3 | 0.567 | 31 | 6.7 |
| Trust in maternal care quality | 12 | 14.8 | 8 | 3.8 | 5 | 2.9 | <0.001 | 9 | 7.0 | 16 | 4.8 | 0.354 | 25 | 5.4 |
| Other barriers |  |  |  |  |  |  |  |  |  |  |  |  |  |  |
| Other | 54 | 66.7 | 127 | 61.1 | 76 | 43.3 | <0.001 | 75 | 58.1 | 182 | 54.3 | 0.40 | 257 | 55.4 |

Factors that were perceived as barriers to giving birth in a healthcare institution at the last childbirth were ordered within the respective accessibility and acceptability theme according to all respondents' affirmative answers. Chi-squared test was used to calculate all *p* values, disregarding missing values.

Overall, 55.4% of the women noted barriers other than the standard response options. Of these, 61.9% also specified other barriers (results not shown), primarily lack of transport and self-motivation. The socio-demographic and SRHR characteristics of the respondents who only answered ‘other’ were similar to the group of women who did not give birth in a healthcare institution. In follow-up qualitative interviews (subsequent study), several women explained that this response reflected a situation where there had been insufficient time to access a healthcare institution because labour had started (results not shown).

### Factors perceived to facilitate the chances of institutional delivery in future pregnancies

The responses from all respondents (*n*=1139), both those who had most recently given birth in a healthcare institution and those who had not, regarding what two factors would be most important for facilitating an institutional delivery in a possible future pregnancy are presented in Table 3. Among accessibility facilitators, companionship emerged as one of the most important factors for just over a third of women in all areas of residence. Transportation and financial resources were also frequently mentioned by women in all areas, particularly in Lunda Sul (transportation, 40.4%; financial resources, 39.9%). Knowing where to go for institutional delivery was chosen as an important facilitator by 23.5% of the women in Huambo and 17.4% in urban areas.

**Table 3.**
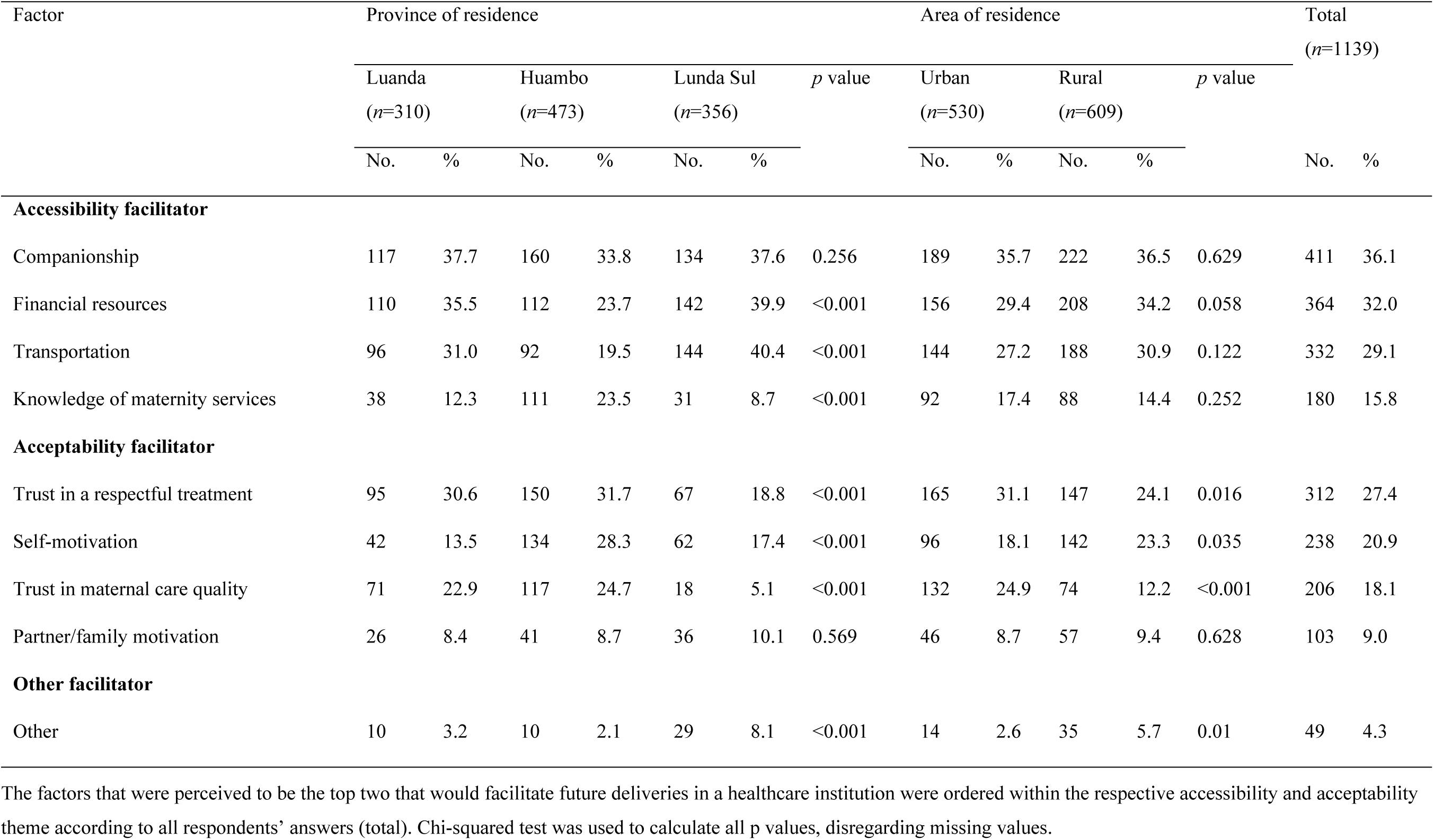
Top two facilitators for future institutional delivery by province and area of residence.

| Factor | Province of residence |  |  |  |  |  |  | Area of residence |  |  |  |  | Total<br>(n=1139) |  |
| --- | --- | --- | --- | --- | --- | --- | --- | --- | --- | --- | --- | --- | --- | --- |
|  | Luanda<br>(n=310) |  | Huambo<br>(n=473) |  | Lunda Sul<br>(n=356) |  | p value | Urban<br>(n=530) |  | Rural<br>(n=609) |  | p value |  |  |
|  | No. | % | No. | % | No. | % |  | No. | % | No. | % |  | No. | % |
| Accessibility facilitator |  |  |  |  |  |  |  |  |  |  |  |  |  |  |
| Companionship | 117 | 37.7 | 160 | 33.8 | 134 | 37.6 | 0.256 | 189 | 35.7 | 222 | 36.5 | 0.629 | 411 | 36.1 |
| Financial resources | 110 | 35.5 | 112 | 23.7 | 142 | 39.9 | <0.001 | 156 | 29.4 | 208 | 34.2 | 0.058 | 364 | 32.0 |
| Transportation | 96 | 31.0 | 92 | 19.5 | 144 | 40.4 | <0.001 | 144 | 27.2 | 188 | 30.9 | 0.122 | 332 | 29.1 |
| Knowledge of maternity services | 38 | 12.3 | 111 | 23.5 | 31 | 8.7 | <0.001 | 92 | 17.4 | 88 | 14.4 | 0.252 | 180 | 15.8 |
| Acceptability facilitator |  |  |  |  |  |  |  |  |  |  |  |  |  |  |
| Trust in a respectful treatment | 95 | 30.6 | 150 | 31.7 | 67 | 18.8 | <0.001 | 165 | 31.1 | 147 | 24.1 | 0.016 | 312 | 27.4 |
| Self-motivation | 42 | 13.5 | 134 | 28.3 | 62 | 17.4 | <0.001 | 96 | 18.1 | 142 | 23.3 | 0.035 | 238 | 20.9 |
| Trust in maternal care quality | 71 | 22.9 | 117 | 24.7 | 18 | 5.1 | <0.001 | 132 | 24.9 | 74 | 12.2 | <0.001 | 206 | 18.1 |
| Partner/family motivation | 26 | 8.4 | 41 | 8.7 | 36 | 10.1 | 0.569 | 46 | 8.7 | 57 | 9.4 | 0.628 | 103 | 9.0 |
| Other facilitator |  |  |  |  |  |  |  |  |  |  |  |  |  |  |
| Other | 10 | 3.2 | 10 | 2.1 | 29 | 8.1 | <0.001 | 14 | 2.6 | 35 | 5.7 | 0.01 | 49 | 4.3 |

The factors that were perceived to be the top two that would facilitate future deliveries in a healthcare institution were ordered within the respective accessibility and acceptability theme according to all respondents' answers (total). Chi-squared test was used to calculate all p values, disregarding missing values.

Among the acceptability facilitators, trust in respectful treatment was selected by 31.7% of the women in Huambo, 30.6% in Luanda and 31.1% in urban areas as one of the two most important factors for future institutional delivery. Self-motivation was also often mentioned by the young women in Huambo and the combined rural areas (28.3% and 23.3%, respectively). Trust in the quality of maternal care was mentioned by 24.7% of the women in Huambo and 22.9% in Luanda, but just 5.1% in Lunda Sul.

### Health literacy characteristics

The characteristics of the respondents’ health literacy (by information sources and SRHR topics) are shown in Table 4 according to whether or not the participants had their last delivery in a healthcare setting. There were significant differences between the two groups for all aspects considered, to the detriment of those who did not give birth in a healthcare institution. For instance, although having received some SRHR information from a health professional was relatively high overall, it was considerably higher among those who had delivered in a healthcare institution (80.6% versus 61.2%); acquiring information on specific SRHR topic areas from more than one source was also higher among those who had delivered in a healthcare institution (see, for example, family planning with 52.9% versus 34.1%).

**Table 4.**
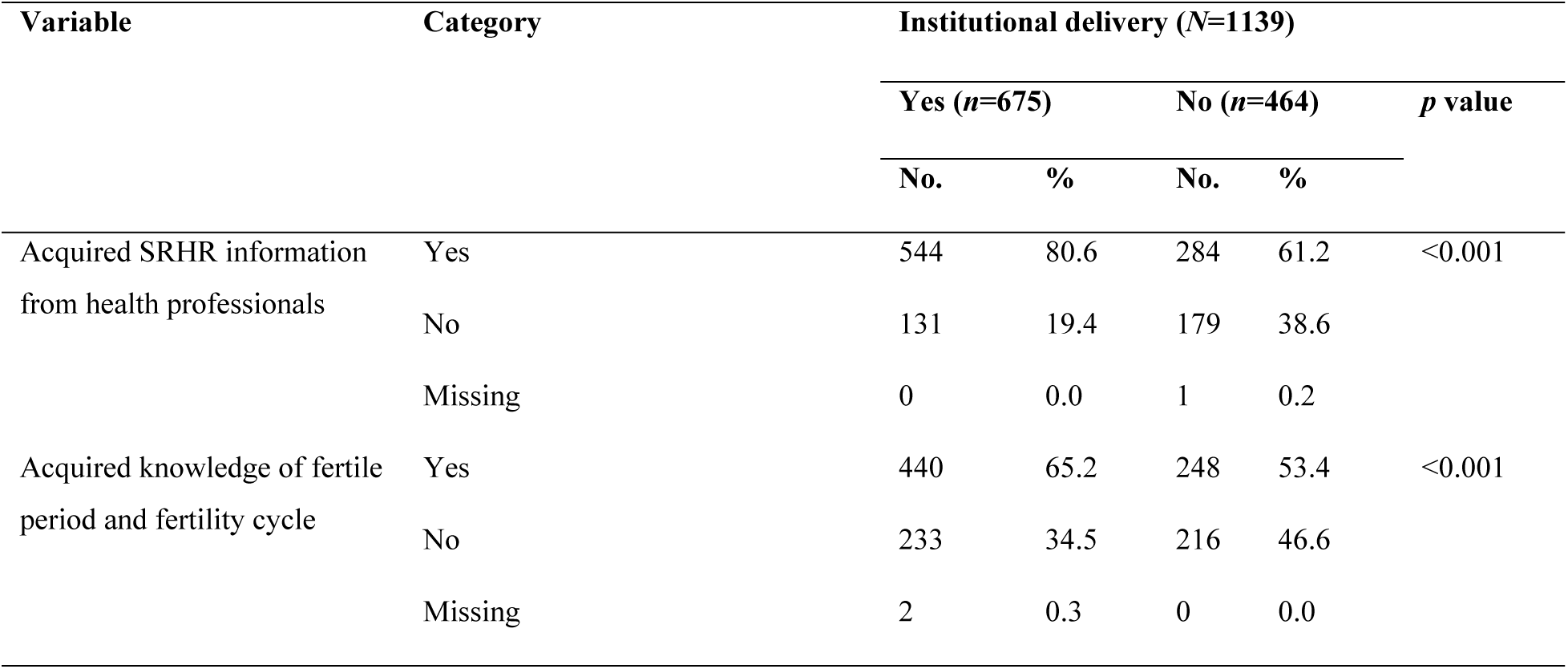
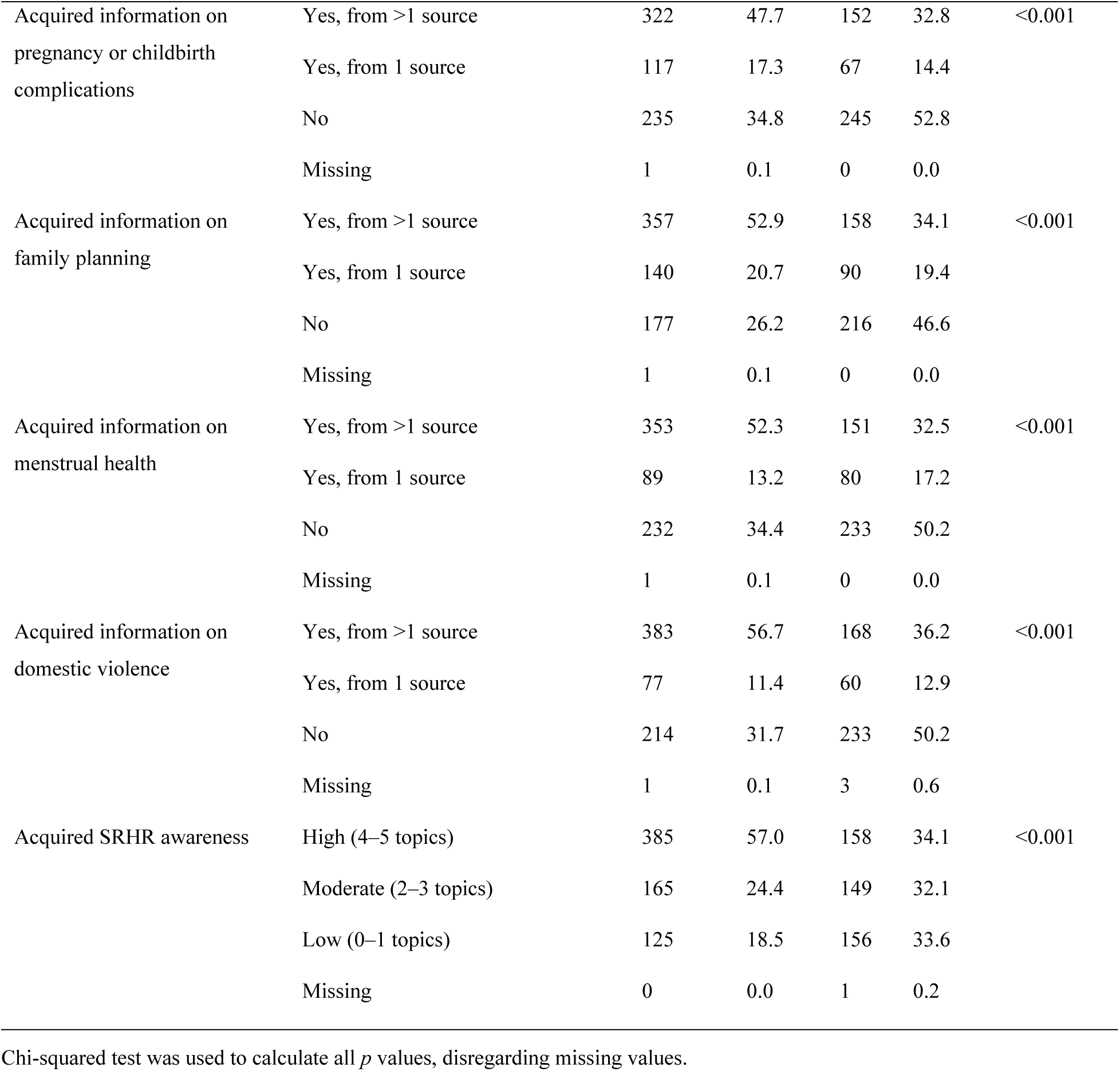
Health literacy characteristics of study participants according to last delivery site.

| Variable | Category | Institutional delivery (N=1139) |  |  |  |  |
| --- | --- | --- | --- | --- | --- | --- |
|  |  | Yes (n=675) |  | No (n=464) |  | p value |
|  |  | No. | % | No. | % |  |
| Acquired SRHR information from health professionals | Yes | 544 | 80.6 | 284 | 61.2 | <0.001 |
|  | No | 131 | 19.4 | 179 | 38.6 |  |
|  | Missing | 0 | 0.0 | 1 | 0.2 |  |
| Acquired knowledge of fertile period and fertility cycle | Yes | 440 | 65.2 | 248 | 53.4 | <0.001 |
|  | No | 233 | 34.5 | 216 | 46.6 |  |
|  | Missing | 2 | 0.3 | 0 | 0.0 |  |
| Acquired information on pregnancy or childbirth complications | Yes, from >1 source | 322 | 47.7 | 152 | 32.8 | <0.001 |
|  | Yes, from 1 source | 117 | 17.3 | 67 | 14.4 |  |
|  | No | 235 | 34.8 | 245 | 52.8 |  |
|  | Missing | 1 | 0.1 | 0 | 0.0 |  |
| Acquired information on family planning | Yes, from >1 source | 357 | 52.9 | 158 | 34.1 | <0.001 |
|  | Yes, from 1 source | 140 | 20.7 | 90 | 19.4 |  |
|  | No | 177 | 26.2 | 216 | 46.6 |  |
|  | Missing | 1 | 0.1 | 0 | 0.0 |  |
| Acquired information on menstrual health | Yes, from >1 source | 353 | 52.3 | 151 | 32.5 | <0.001 |
|  | Yes, from 1 source | 89 | 13.2 | 80 | 17.2 |  |
|  | No | 232 | 34.4 | 233 | 50.2 |  |
|  | Missing | 1 | 0.1 | 0 | 0.0 |  |
| Acquired information on domestic violence | Yes, from >1 source | 383 | 56.7 | 168 | 36.2 | <0.001 |
|  | Yes, from 1 source | 77 | 11.4 | 60 | 12.9 |  |
|  | No | 214 | 31.7 | 233 | 50.2 |  |
|  | Missing | 1 | 0.1 | 3 | 0.6 |  |
| Acquired SRHR awareness | High (4–5 topics) | 385 | 57.0 | 158 | 34.1 | <0.001 |
|  | Moderate (2–3 topics) | 165 | 24.4 | 149 | 32.1 |  |
|  | Low (0–1 topics) | 125 | 18.5 | 156 | 33.6 |  |
|  | Missing | 0 | 0.0 | 1 | 0.2 |  |

Chi-squared test was used to calculate all *p* values, disregarding missing values.

Fewer women in both groups had acquired information on complications related to pregnancy or childbirth from one (17.3% versus 14.4%) or more than one source (47.7% versus 32.8%). More of those who had given birth in a healthcare institution had acquired a high level of SRHR awareness (57%) compared with those who had had non-institution delivery (34.1%).

### Associations between health literacy and institutional delivery

Associations between health literacy attributes and institutional delivery are presented in Table 5. Significant associations were found in two of the three provinces (Luanda and Huambo) and in both types of residential areas (urban and rural). However, the adjusted odds ratios (aORs) were more pronounced at the province level, especially with regard to having health personnel as an SRHR information source (Luanda: aOR, 2.74; 95% confidence interval [CI], 1.32–5.67; Huambo: aOR, 2.44; 95% CI, 1.39–4.28; urban area: aOR, 2.02; 95% CI, 1.14–3.59; rural area: aOR, 1.58; 95% CI, 1.09–2.29).

**Table 5.** Associations between health literacy and institutional delivery stratified by province and area of residence.

| Variable | Category | Province of residence, aOR (95% CI) |  |  | Area of residence, aOR (95% CI) |  | Total, aOR (95% CI)<br>(n=1139) |
| --- | --- | --- | --- | --- | --- | --- | --- |
|  |  | Luanda (n=310) | Huambo (n=473) | Lunda Sul (n=356) | Urban (n=530) | Rural (n=609) |  |
| Acquired SRHR information from health professional | Yes | 2.74 (1.32–5.67) | 2.44 (1.39–4.28) | 1.09 (0.69–1.73) | 2.02 (1.14–3.59) | 1.58 (1.09–2.29) | 1.58 (1.17–2.13) |
|  | No | 1 | 1 | 1 | 1 | 1 | 1 |
| Acquired knowledge of fertile period and cycle | Yes | 2.09 (1.16–3.78) | 0.89 (0.56–1.42) | 1.13 (0.68–1.87) | 1.63 (1.02–2.59) | 1.01 (0.69–1.46) | 1.13 (0.86–1.48) |
|  | No | 1 | 1 | 1 | 1 | 1 | 1 |
| Acquired information on pregnancy or childbirth complications | Yes, from >1 source | 3.86 (1.92–7.77) | 1.54 (0.95–2.50) | 1.06 (0.58–1.90) | 2.12 (1.28–3.50) | 1.43 (0.96–2.14) | 1.52 (1.14–2.04) |
|  | Yes, from 1 source | 1.88 (0.91–3.90) | 1.78 (0.89–3.58) | 1.11 (0.58–2.15) | 2.22 (1.16–4.24) | 1.03 (0.61–1.72) | 1.35 (0.91–2.04) |
|  | No | 1 | 1 | 1 | 1 | 1 | 1 |
| Acquired information on family planning | Yes, from >1 source | 2.57 (1.26–5.27) | 2.24 (1.33–3.77) | 0.90 (0.50–1.64) | 2.00 (1.13–3.54) | 1.59 (1.04–2.41) | 1.62 (1.19–2.20) |
|  | Yes, from 1 source | 1.66 (0.74–3.75) | 1.82 (0.96–3.44) | 1.12 (0.62–2.03) | 1.31 (0.69–2.46) | 1.42 (0.90–2.41) | 1.28 (0.89–1.85) |
|  | No | 1 | 1 | 1 | 1 | 1 | 1 |
| Acquired information on menstrual health | Yes, from >1 source | 2.41 (1.23–4.69) | 1.72 (1.04–2.83) | 0.99 (0.53–1.82) | 2.28 (1.37–3.82) | 1.36 (0.90–2.06) | 1.50 (1.12–2.02) |
|  | Yes, from 1 source | 1.48 (0.63–3.49) | 0.80 (0.42–1.54) | 0.89 (0.45–1.75) | 1.59 (0.84–3.03) | 0.67 (0.39–1.14) | 0.89 (0.60–1.31) |
|  | No | 1 | 1 | 1 | 1 | 1 | 1 |
| Acquired information on domestic violence | Yes, from >1 source | 2.12 (1.06–4.23) | 1.11 (0.68–1.81) | 1.06 (0.59–1.90) | 1.47 (0.88–2.48) | 1.26 (0.84–1.90) | 1.30 (0.95–1.77) |
|  | Yes, from 1 source | 1.25 (0.47–3.32) | 0.82 (0.43–1.57) | 1.71 (0.82–3.56) | 1.57 (0.71–3.49) | 0.94 (0.56–1.57) | 1.06 (0.70–1.61) |
|  | No | 1 | 1 | 1 | 1 | 1 | 1 |
| Acquired SRHR awareness | High (4–5 topics) | 3.99 (1.72–9.23) | 2.11 (1.08–4.12) | 1.05 (0.54–2.04) | 2.87 (1.49–5.52) | 1.48 (0.91–2.39) | 1.57 (1.11–2.23) |
|  | Moderate (2–3 topics) | 1.69 (0.72–3.97) | 1.46 (0.72–2.96) | 0.90 (0.54–1.51) | 1.71 (0.87–3.37) | 0.92 (0.60–1.41) | 0.98 (0.69–1.40) |
|  | Low (0–1 topics) | 1 | 1 | 1 | 1 | 1 | 1 |

The association between health literacy and institutional delivery was particularly strong among women from Luanda. When the yes category was split into one versus more than one source, the odds tended to be significantly higher with more than one source, and more often in Luanda and in urban areas. For instance, having acquired information on domestic violence from more than one source was only significantly associated with giving birth in a healthcare institution among the women in Luanda (aOR, 2.12; 95% CI, 1.06–4.23). In addition, when information about pregnancy or childbirth complications was acquired from more than one source as opposed to not acquiring this information (0 source), the odds of giving birth in a healthcare institution were almost four times as high for the women in Luanda (aOR, 3.86; 95% CI, 1.92–7.77) and more than double for the women in urban areas (aOR, 2.12; 95% CI, 1.28– 3.50).

In addition, in comparison with those who had no information about family planning, the women who had more than one source of SRHR information, the odds of giving birth in a healthcare institution were twice as high in Luanda (aOR, 2.57; 95% CI, 1.26–5.27), Huambo (aOR, 2.24; 95% CI, 1.33–3.77) and urban areas (aOR, 2.00; 95% CI, 1.13–3.54). In the same geographic areas, high SRHR awareness also increased the odds of having had an institutional delivery (Luanda: aOR, 3.99; 95% CI, 1.72–9.23; urban areas: aOR, 2.87; 95% CI, 1.49–5.52; Huambo: aOR, 2.11; 95% CI, 1.08–4.12).

## Discussion

### Main findings

The study showed that six out of ten had undergone institutional delivery. Compared with those who did not, those who did were more likely to reside in urban areas, be more educated, and wealthier.

The most common barrier put forward by those who had not given birth in a healthcare institution at their last birth (40.7%) was, by far, lack of transport, followed by a lack of self-motivation and, thereafter, financial constraints. In contrast, all respondents aggregated, when asked about factors that would make it easier to give birth in such an institution in the future, the ranking changed remarkably, starting with being accompanied, closely followed by financial resources, transport and trust in a respectful treatment.

Although not having information on fundamental SRHR topics was common among all respondents, this was particularly salient among those who had not given birth in a healthcare institution.

In all areas of residence and provinces except in Lunda Sul, the province with the most challenging living conditions, there were significant associations between the health literacy measures and place of delivery. Rather than being graded, significant associations arose at the highest levels of SRHR literacy, most remarkably for those who had acquired SRHR information from several sources (versus 0 or 1 source) and who had a high degree (versus a low or medium degree) of SRHR awareness.

### Young women’s perceptions on the conditions for institutional delivery

The present study is coherent with previous findings regarding maternal healthcare accessibility in Angola and similar settings. For instance, the proportions of institutional births (59.2%) and births overseen by health professionals (57.1%) were largely consistent with official statistics on Angola (54.2% and 60.0%, respectively) [61]. Likewise, in spite of differences in the study groups and areas, research from other sub-Saharan put forward similar types of factors relative to institutional delivery with transport infrastructure and household wealth levels being particularly highlighted as influencing the use of institutional versus home delivery [2,6,7,12–17]. Interestingly, the study indicated a form of environmental reliance that has received comparatively little attention in previous studies: the respondents’ prioritisation of companionship as a key factor for the realisation of institutional delivery [3,8]. Whether this is more specific to younger women remains to be determined, but the findings underscore the multifaceted nature and the inherently vulnerable circumstances of childbirth for a young woman. Eventually, it might be essential to conceive promotion strategies incorporating the provision of emotional support.

The intricacy of the issue is also reflected in previous research findings showing that the substantial familial and domestic responsibilities of women residing in low-income regions impede women from seeking maternity services [12]. The fact that many participants, particularly from Huambo, identified self-motivation as a key factor, despite the vast majority designating a healthcare institution as the optimal place to give birth, can be interpreted in relation to such research findings. For many women, utilising institutional delivery services involves a negotiation between this and the wider context of life, encompassing the family’s needs and the financial implications of any out-of-home childbirth [12]. The study also reveals several other types of precarious factors to take into consideration, including e.g., IPV and severe food insecurity, as these have the potential to diminish resilience and the motivation for institutional childbirth. When interpreting the phenomenon of young women’s self-motivation, it is therefore essential to reflect on its interplay with the overall life situation and perceptions of their realistic options for institutional delivery. That is to say, self-motivation may be an indication of what women consider feasible rather than personal preferences only.

The study lends further support to the notion that socio-economic characteristics of high-risk groups should be given due consideration for the augmentation of institutional delivery [1–3,5–8,10–16]. It also provides a basis for reflecting upon the notion that successful interventions in this domain may require the incorporation of strategies to address emotional support needs as well as an insightful understanding of the dynamics of women’s self-motivation [2,14].

### The potential of health literacy to promote young women’s health in Angola

Previous research has shown that goals other than those related to a specific health topic may well compete with and undermine an individual’s prioritization of health-promoting behaviour even when adequate health knowledge is available [22,25,30,33]. Correspondingly, it is imperative to recognize that increasing young women’s health literacy alone is insufficient to resolve the complex challenges to their SRHR.

A specific commentary on the issue of the social gradient of health literacy is warranted. This is well documented and also came across in the current study, for example in that associations between health literacy and institutional delivery pertained primarily to women residing in the wealthier living areas, such as the capital province or urban areas. Notably, in Lunda Sul, the province with the greatest socio-economic challenges [57,61,68–71], no links between health literacy and childbirth location were identified, irrespective of the level of health literacy of the participants. The study is silent regarding the reasons behind variations related to living area. Nevertheless, socio-economic factors such as educational level and income would be in line with previous research on social determinants of institutional delivery [6,7,12–16,42,43] as well as health literacy [25,30,33,36,39,42].

Notwithstanding the many challenges faced by young Angolan women, the study demonstrates that there are circumstances where health literacy might help promoting desired SRHR outcomes like giving birth in a healthcare institution. Except for one province, the most precarious one, associations between health literacy and childbirth setting were common in the other living areas. This is particularly encouraging given that this applied to SRHR topics that do not require a high educational level or access to specific types of information media for full understanding. Instead, the topics were of a fundamental nature and arguably indispensable for a young woman’s comprehension of her body and the means to preserve it optimally, both autonomously and in interaction with others [10,11,29,80].

In addition, the associations found were not exclusive to SRHR topics closely related to the delivery situation but also encompassed more general topics such as family planning and menstrual health. Additionally, the provision of SRHR information by health professionals was favourable, irrespective of the specific SRHR subjects addressed in the exchange. It showed, likewise, important to acquire information on a wide range of SRHR topics, once again irrespective of the specific SRHR topics included. The fact that these more general aspects of health information acquisition and exchange were found to be beneficial point to that a more generic health literacy may also have an empowering and health promotion potential [28,29].

That significant associations appear for categories indicative of higher levels of health literacy is a result of high interest. It underscores the literature’s emphasis on the progression towards higher and more comprehensive health literacy levels, with the more advanced levels being more effective in promoting health [23–27]. Access to information alone was insufficient; a broad range of information sources or SRHR topics was essential to establish a sufficiently robust health literacy base. Consequently, the utilisation of a variety of sources for conveying information, emphasising different aspects of the respective SRHR issue, could be pivotal in ensuring its comprehensive uptake. This insight reinforces the notion of broadly targeted interventions [7,40,41,45,49,50], incorporating the wider environment of young women.

The importance of involving health professionals in such interventions is emphasised in light of the tangible impact the acquisition of SRHR information from such a source had. In addition, other research has highlighted the health sector as a key player in promoting women’s SRHR knowledge, particularly in terms of the extent to which health professionals take responsibility for providing information on key issues and the manner in which they ensure its reception by women from diverse educational backgrounds [7,19,32,35,37,40]. It is noteworthy in itself that so few women reported having received information on complications associated with pregnancy or childbirth (approximately half or two-thirds, depending on place of delivery), but even more so given the predominance of antenatal attendance during pregnancy among the surveyed population (surpassing 80%, irrespective of the subsequent delivery location), where such information could have been provided. This resonates with research that has shown an association between the quality of antenatal care with healthcare institution birth [6,7,11,16,42]. It underscores the necessity to fortify the capacities and roles of health professionals, particularly in increasing institutional childbirth access for young women.

The pervasive disparities in health literacy, in conjunction with the environmental factors identified by women as impediments to institutional delivery, substantiate assertions that multisectoral action is imperative to address the underlying reasons for out-of-hospital childbirths and other social determinants of maternal mortality [1–3,5,8,21,62–66]. Thus, health literacy initiatives should be implemented in conjunction with other long-term efforts to enhance access to quality healthcare services. These initiatives ought to be underpinned by comprehensive integration of gender-sensitive and social determinant perspectives in interventions [22,27,32–37,43–45,49,60], thereby ensuring a multifaceted and sustained approach to addressing the prevailing problems.

### Strength and limitations

A notable strength of the study is the high response rate as well as the size and diversity of the context of the young participants. By introducing variability in their living areas and conditions, it was possible to get a sense of whether and how the prevalence of giving birth in a healthcare institution and related factors varied. A wide range of health literacy variables was included, such as different SRHR topics and sources of information. Furthermore, the composition of the questions facilitated analysis of health literacy levels, thereby presenting an original contribution to the health literacy field. The study’s approach enabled examination of the significance of health literacy comprehensiveness, which, to the best of our knowledge, has not been undertaken previously. The large number of individual variables also made it possible to control for several socio-economic factors presumed to be linked to health service provision.

In addition, this work was meticulously grounded in robust theoretical literature, as described above, and its application in the specific context was preceded by thorough piloting with the target group to ensure relevance and appropriateness of the questions. For example, it was recognized that the context precludes the assumption of the indispensability of formal education and literacy as components of health literacy, as frequently presumed in existing research [e.g. 25]. The health literacy survey questions constructed for the present study were crafted to encompass health information acquired, irrespective of its mode of transmission, and adjustments were made for literacy and other socio-economic factors.

The quality of the data ensured by training the data collectors and regular quality control contribute to the quality and validity of the data. In light of the high degree of illiteracy and limited experience with questionnaire-based methods among a substantial proportion of the study population, the research team decided to use an interview-based method for data collection as opposed to a self-administered approach. Several measures were implemented to establish an interviewing environment that was both comfortable and receptive. These measures were driven by ethical considerations, as well as the desire to mitigate any desirability bias that may arise from a potential power imbalance between the interviewer and participant. Moreover, the careful design and subsequent adaptation to the local context of the survey aimed to reduce the risk of misclassification.

Notwithstanding, the study has identified areas for further research. A notable proportion of the respondents indicated barriers to institutional delivery that were not identified during the development of the questionnaire and, therefore, were not included in the standardized response options. This finding indicates the existence of barriers not highlighted in previous research or anticipated by participants in the pilot test of the questionnaire. The response options related to access to institutional delivery could therefore benefit from further exploration, preferably using qualitative in-depth interviews.

Further research is also required into what mechanisms and processes lead to levels of health literacy that have been shown to promote health. In addition, there is a need for a more in-depth exploration of the interplay between individual, community and health professionals’ health literacy in ensuring that young women’s access to institutional delivery. A review of the SRHR content from the respective information sources and an investigation into young women’s trust in different information providers would also be a valuable addition to this field. This was not covered in the present study, yet it would further contribute to the health literacy field and support the evidence-based development of interventions to strengthen women’s health and rights.

The areas that have been identified for further research should be considered when interpreting the findings. Furthermore, the study’s focus on a particular age group, and its utilization of a non-random sample, are aspects that demand careful consideration. These factors preclude any claims of the results being generalizable to the country as a whole or to Angolan women in general. Nevertheless, the study shows reasonable consistency with other studies with respect to the socio-economic characteristics of young women [52,56–58,61], and it mirrors official statistics in terms of the considerable variation between the provinces included [57,61,72]. Consequently, it can be reasonably assumed that the respondents share many life circumstances with young women in the country and other similar contexts; therefore, our findings have the potential to provide valuable insights into issues related to health literacy and institutional delivery access in comparable populations.

Finally, a limitation of this study is the cross-sectional design, which does not allow for causal effects. For example, it cannot be determined whether women delivered in a healthcare institution because they had information about pregnancy or childbirth complications or whether they had this information because they had given birth in a healthcare institution.

## Conclusion

The study draws attention to the need to increase the likelihood of young Angolan women delivering in healthcare institutions. Its findings suggest that interventions in this realm could usefully include innovative, cost-effective solutions that allow for advanced planning; the establishment of transportation arrangements; and the support of family members during labour [3,5,8].

In addition, the study supports the view that health literacy has the potential to promote the SRHR of young women, also in the context of restricted access to education, basic livelihoods and maternal healthcare. The identification of various associations between health literacy and institutional delivery highlights the importance of incorporating knowledge-enhancing efforts into interventions aiming to strengthen young women’s SRHR.

Collaborative efforts ought to embrace the socio-economic and institutional milieu in which young women formulate their decisions and apply their knowledge [6–8]. Without this, the effectiveness of health promotion interventions is anticipated to be diminished and their capacity to address health inequalities may be compromised [1,5,8,25].

## Data Availability

Data are fully available without restriction.

## References

1. Starrs AM, Ezeh AC, Barker G, Basu A, Bertrand JT, Blum R, et al. Accelerate progress— sexual and reproductive health and rights for all: report of the Guttmacher–Lancet Commission. Lancet. 2018; 391(10140): 2642–92.

2. Doctor HV, Nkhana-Salimu S, Abdulsalam-Anibilowo M. Health facility delivery in sub-aharan Africa: successes, challenges, and implications for the 2030 development agenda. BMC Public Health. 2018; 18(1): 765.

3. Moyer CA, Lawrence ER, Beyuo TK, Tuuli MG, Oppong SA. Stalled progress in reducing maternal mortality globally: what next? Lancet. 2023; 401(10382): 1060–2.

4. World Health Organization (WHO). Trends in maternal mortality 2000 to 2020: estimates by WHO, UNICEF, UNFPA, World Bank Group and UNDESA/Population Division. 2023. Available from: https://www.who.int/publications-detail-redirect/9789240068759.

5. World Health Organization (WHO), United Nations Population Fund (UNFPA). Ending preventable maternal mortality (EPMM). A renewed focus for improving maternal and newborn health and wellbeing. 2021. Available from: https://cdn.who.int/media/docs/default-source/mca-documents/maternal-nb/ending-preventable-maternal-mortality_epmm_brief-230921.pdf?sfvrsn=f5dcf35e_5.

6. Olubodun T, Rahman SA, Odukoya OO, Okafor IP, Balogun MR. Determinants of health facility delivery among young mothers aged 15-24 years in Nigeria: a multilevel analysis of the 2018 Nigeria demographic and health survey. BMC Pregnancy Childbirth. 2023; 23(1): 185.

7. Nyamtema AS, Mwakatundu N, Dominico S, Mohamed H, Pemba S, Rumanyika R, et al. Enhancing maternal and perinatal health in under-served remote areas in sub-Saharan Africa: a Tanzanian model. PLoS One. 2016; 11(3): e0151419.

8. Campbell OMR, Graham WJ. Strategies for reducing maternal mortality: getting on with what works. Lancet. 2006; 368(9543): 1284–99.

9. World Health Organization (WHO). The Global Health Observatory. Explore a world of health data. Indicator Meta Data Register List: Proportion of births delivered in a health facility (Facility births) (%). 2025. Available from: https://www.who.int/data/gho/indicator-metadata-registry/imr-details/institutional-birth.

10. African Commission on Human and People’s Rights (ACHPR). General Comment No. 2 on Article 14.1 (a), (b), (c) and (f) and Article 14.2 (a) and (c) of the Protocol to the African Charter on Human and Peoples’ Rights on the Rights of Women in Africa. 2014. Available from: https://vcsafund.org/wp-content/uploads/2024/09/achprinstrgeneralcomment2rightsofwomeninafricaengpdf.pdf.

11. United Nations Committee on Economic, Social and Cultural Rights (UN ESCR). General Comment No. 22 (2016) on the Right to Sexual and Reproductive Health (Article 12 of the International Covenant on Economic, Social, and Cultural Rights). Available from: https://digitallibrary.un.org/record/832961/files/E_C.12_GC_22-EN.pdf?ln=en.

12. Bain LE, Aboagye RG, Dowou RK, Kongnyuy EJ, Memiah P, Amu H. Prevalence and determinants of maternal healthcare utilisation among young women in sub-Saharan Africa: cross-sectional analyses of demographic and health survey data. BMC Public Health. 2022; 22(1): 647.

13. Dahlab R, Sakellariou D. Barriers to accessing maternal care in low income countries in Africa: a systematic review. Int J Environ Res Public Health. 2020; 17(12): 4292.

14. Kota K, Chomienne MH, Geneau R, Yaya S. Socio-economic and cultural factors associated with the utilization of maternal healthcare services in Togo: a cross-sectional study. Reprod Health. 2023; 20(1): 109.

15. Rosário EVN, Gomes MC, Brito M, Costa, D. Determinants of maternal health care and birth outcome in the Dande Health and Demographic Surveillance System area, Angola. PLoS One. 2019; 14(8): e0221280.

16. Samuel O, Zewotir T, North D. Decomposing the urban–rural inequalities in the utilisation of maternal health care services: evidence from 27 selected countries in Sub-Saharan Africa. Reprod Health. 2021; 18(1): 1–216.

17. Aoki A, Mochida K, Kuramata M, Sadamori T, Sapalalo P, Tchicondingosse L, et al. Association between the quality of care and continuous maternal and child health service utilisation in Angola: longitudinal data analysis. J Glob Health. 2023; 13: 04073.

18. Rosen HE, Lynam PF, Carr C, Reis V, Ricca J, Bazant ES, et al. Quality of maternal and newborn care study group of the maternal and child health integrated program. Direct observation of respectful maternity care in five countries: a cross-sectional study of health facilities in East and Southern Africa. BMC Pregnancy Childbirth. 2015; 15: 306.

19. World Health Organization (WHO). The prevention and elimination of disrespect and abuse during facility-based childbirth: WHO statement. 2014. Available from: https://iris.who.int/bitstream/handle/10665/134588/WHO_RHR_14.23_eng.pdf?sequence=1.

20. Ansu-Mensah M, Danquah FI, Bawontuo V, Ansu-Mensah P, Kuupiel D. Maternal perceptions of the quality of care in the free maternal care policy in sub-Saharan Africa: a systematic scoping review. BMC Health Serv Res. 2020; 20(1): 911.

21. Pavalagantharajah S, Negrin AR, Bouzanis K, Joan Lee TS, Miller P, Jones R, et al. Facility-based maternal quality of care frameworks: a systematic review and best fit framework analysis. Matern Child Health J. 2023; 27(10): 1742–53.

22. Nutbeam D. Health literacy as a public health goal: a challenge for contemporary health education and communication strategies into the 21st century. Health Promot Int. 2000; 15(3): 259–68.

23. Peerson A, Saunders M. Health literacy revisited: what do we mean and why does it matter?. Health Promot Int. 2009; 24(3): 285–96.

24. Paasche-Orlow M, Wolf MS. The causal pathways linking health literacy to health outcomes. Am J Health Behav. 2007; 31(Suppl): S19–26.

25. Nutbeam D, Lloyd JE. Understanding and responding to health literacy as a social determinant of health. Annu Rev Public Health. 2021; 42:159–73.

26. Sykes S, Wills J, Rowlands G, Popple K. Understanding critical health literacy: a concept analysis. BMC Public Health. 2013; 13: 150.

27. Sørensen K, Van den Broucke S, Fullam J, Doyle G, Pelikan J, Slonska Z, et al. Health literacy and public health: a systematic review and integration of definitions and models. BMC Public Health. 2012; 12(1): 80.

28. Mårtensson L, Hensing G. Health literacy - a heterogeneous phenomenon: a literature review. Scand J Caring Sci. 2012; 26: 151–60.

29. Kickbusch I, Wait S, Maag D, McGuire P, Banks I. Navigating health: the role of health literacy. Alliance for Health and the Future, International Longevity Centre-UK. 2006. Available from: https://ilcuk.org.uk/wp-content/uploads/2018/10/NavigatingHealth.pdf.

30. Dolezel D, Hewitt B. Social determinants of health literacy: a cross-sectional exploratory study. Health Promot Int. 2023; 38(5): daad127.

31. Berkman ND, Sheridan SL, Donahue KE, Halpern DJ, Crotty KP. Low health literacy and health outcomes: an updated systematic review. Ann Intern Med. 2011; 155(2): 97–107.

32. Kilfoyle KA, Vitko M, O’Conor R, Bailey SC. Health literacy and women’s reproductive health: a systematic review. J Womens Health (Larchmt). 2016; 25(12): 1237–55.

33. Stormacq C, Van den Broucke S, Wosinski J. Does health literacy mediate the relationship between socioeconomic status and health disparities? Integrative review. Health Promot Int. 2019; 34: e1–17.

34. Renkert S, Nutbeam D. Opportunities to improve maternal health literacy through antenatal education: an exploratory study. Health Promot Int. 2001; 16(4): 381–8.

35. Nawabi F, Krebs F, Vennedey V, Shukri A, Lorenz L, Stock S. Health literacy in pregnant women: a systematic review. Int J Environ Res Public Health. 2021; 18(7): 3847.

36. Holmen H, Flølo T, Tørris C, Løyland B, Almendingen K, Bjørnnes AK, et al. Unpacking the public health triad of social inequality in health, health literacy, and quality of life—a scoping review of research characteristics. Int J Environ Res Public Health. 2024; 21(1): 36.

37. Baumeister A, Chakraverty D, Aldin A, Seven ÜS, Skoetz N, Kalbe E, et al. The system has to be health literate, too” - perspectives among healthcare professionals on health literacy in transcultural treatment settings. BMC Health Serv Res. 2021; 21: 716.

38. Paucar-Caceres A, Vílchez-Román C, Quispe-Prieto S. Health literacy concepts, themes, and research trends globally and in Latin America and the Caribbean: a bibliometric review. Int J Environ Res Public Health. 2023; 20(22): 7084.

39. Amanu, A, Birhanu Z, Godesso A. Sexual and reproductive health literacy among young people in Sub-Saharan Africa: evidence synthesis and implications. Global Health Action. 2023; 16(1): 2279841.

40. Trezona A, Dodson S, Osborne RH. Development of the organisational health literacy responsiveness (Org-HLR) framework in collaboration with health and social services professionals. BMC Health Serv Res. 2017; 17(1): 513.

41. Koh HK, Baur C, Brach C, Harris LM, Rowden JN. Toward a systems approach to health literacy research. J Health Commun. 2013; 18(1): 1–5.

42. Bello CB, Esan DT, Akerele SA, Fadare RI. Maternal health literacy, utilisation of maternal healthcare services and pregnancy outcomes among newly delivered mothers: a cross-sectional study in Nigeria. Public Health Pract (Oxf). 2022; 3: 100266.

43. Dankwah E, Zeng W, Feng C, Kirychuk S, Farag M. The social determinants of health facility delivery in Ghana. Reprod Health. 2019; 16(1): 101.

44. Barchi F, Ntshebe O, Apps H, Ramaphane P. Contraceptive literacy among school-going adolescents in Botswana. Int Nurs Rev. 2022; 69(1): 86–95.

45. Heine M, Lategan F, Erasmus M, Lombaard CM, McCarthy N, Olivier J, et al. Health education interventions to promote health literacy in adults with selected non-communicable diseases living in low-to-middle income countries: a systematic review and meta-analysis. J Eval Clin Pract. 2021; 27: 1417–28.

46. Pelikan JM, Ganahl K, Roethlin F. Health literacy as a determinant, mediator and/or moderator of health: empirical models using the European Health Literacy Survey dataset. Glob Health Promot. 2018; 25(4): 57–66.

47. World Health Organization (WHO). Health literacy development for the prevention and control of noncommunicable diseases: volume 1: overview 2022. World Health Organization: Geneva, Switzerland. Available from: https://iris.who.int/bitstream/handle/10665/364204/9789240055353-eng.pdf?sequence=1.

48. Nutbeam D. Improving health literacy: how to succeed. Public Health Res Pract. 2023; 33(1): e3312301.

49. Sharma A, Ronan A, Namiba A, Oktariani A, Davies L. Beyond toolkits: sexual and reproductive health and rights literacy requires women-centred structures, services and policies. J Int AIDS Soc. 2022; 25(3): e25888.

50. Chipako I, Singhal S, Hollingsworth B. Impact of sexual and reproductive health interventions among young people in sub-Saharan Africa: a scoping review. Front Glob Womens Health. 2024; 5: 1344135.

51. World Health Organization (WHO). Operational framework for monitoring social determinants of health equity. Geneva: World Health Organization; 2024. Available from: https://iris.who.int/bitstream/handle/10665/375732/9789240088320-eng.pdf?sequence=1.

52. Global Change Data Lab (GCDL). Our world in data: Angola. Available from: https://globaldatalab.org/areadata/profiles/AGOt/.

53. Mosaiko Relatório da pesquisa sobre políticas públicas inclusivas numa perspectiva de género, 2019-2021, Luanda: Tipografia Coimbra. 2021. Available from: https://mosaiko.op.org/wp-content/uploads/2019/05/PAPPIA-Relatorio-de-Pesquisa-Web.pdf.

54. Strønen IÅ, Nangacovie M. The gendering of poverty and inequality in rural Malanje, Angola. CMI Report R 2018:10. Bergen: Chr. Michelsen Institute. Available from: https://www.cmi.no/publications/6567-the-gendering-of-poverty-and-inequality-in-rural.

55. Koerner E, Zandamela J, Duckett A. Menstrual management in Angola: effectiveness of providing quality menstrual products and educational workshops in Huambo, Huíla, Luanda and Lunda Sul. UNFPA Learning Study. 2021. Available from: https://angola.unfpa.org/sites/default/files/pub-pdf/ebe_girl_unfpa_angola_mh_study_phase_1_en_2021-05-24_final.pdf.

56. World Health Organization (WHO). Global Health Observatory data repository. Maternal and reproductive health. Available from: https://apps.who.int/gho/data/node.main.SRHIB?lang=en.

57. Instituto Nacional de Estatística (INE), Minstério da Saúde (MINSA), Ministério do Planeamento e do Desenvolvimento Territorial (MINPLAN), ICF. Angola inquérito de indicadores múltiplos e de saúde (IIMS) 2015-2016. 2017. Available from: https://www.dhsprogram.com/pubs/pdf/SR238/SR238.P.pdf.

58. Pacatolo C, Boio D, Kitombe C. Angolans dissatisfied with government efforts to promote equal rights for women. Afrobarometer Dispatch No. 622, 28 March 2023. Available from: https://www.afrobarometer.org/wp-content/uploads/2023/03/AD622-Angolans-dissatisfied-with-government-performance-on-gender-equality-Afrobarometer-28march2023.pdf.

59. Sama CB, Ngasa SN, Dzekem BS, Choukem SP. Prevalence, predictors and adverse outcomes of adolescent pregnancy in sub-Saharan Africa: a protocol of a systematic review. Syst Rev. 2017; 6(1): 247.

60. Asmamaw DB, Tafere TZ, Negash WD. Prevalence of teenage pregnancy and its associated factors in high fertility sub-Saharan Africa countries: a multilevel analysis. BMC Women’s Health. 2023; 23(1): 23.

61. Instituto Nacional de Estatística (INE), Minstério da Saúde (MINSA), Inner City Fund (ICF). 2024. Inquérito de indicadores múltiplos e de saúde de Angola, 2023–2024: relatório de indicadores básicos. Available from: https://www.ine.gov.ao/Arquivos/arquivosCarregados/Carregados/Publicacao_638487571879652908.pdf.

62. Umar AS, Kabamba L. Maternal mortality in the main referral hospital in Angola, 2010-2014: understanding the context for maternal deaths amidst poor documentation. Int J MCH AIDS. 2016; 5(1): 61–71.

63. de Almeida N, Teixeira A, Capoco Sachiteque A, Molina JR, dos Prazeres Tavares H, et al. Characterisation of induced abortion and consequences to women’s health at Hospital Central do Huambo – Angola. J Obstet Gynecol. 2019; 40(4): 558–63.

64. Oliveira D, de Oliveira JM, Martins MdR, Barosso MR, Castro R, Cordeiro L, et al. Maternal profiles and pregnancy outcomes: a descriptive cross-sectional study from Angola. Matern Child Health J. 2023; 27: 2091–8.

65. Prata N, Bell S, Fraser A, Carvalho A, Neves I, Nieto-Andrade B. Partner support for family planning and modern contraceptive use in Luanda, Angola. Afr J Reprod Health. 2017; 21(2): 35–48.

66. Yaya S, Ghose B. Prevalence of unmet need for contraception and its association with unwanted pregnancy among married women in Angola. PLoS One. 2018; 13(12): e0209801.

67. United States Agency for International Development (USAID). The Demographic and Health Survey program. DHS model questionnaires 2014, adapted for Angola. Available from: https://dhsprogram.com/Methodology/Survey-Types/DHS-Questionnaires.cfm#CP_JUMP_16179.

68. Leander S, Alexandre SB, Vinha KP, Monsalve Montiel EM, Nguyen MC, Medina Giopp A, et al. Angola poverty assessment (English). Washington, DC: World Bank Group. 2020. Available from: http://documents.worldbank.org/curated/en/328741593674436204/Angola-Poverty-Assessment.

69. Rodrigues CU, Bryceson DF. Precarity in Angolan diamond mining towns, 1920–2014: tracing agency of the state, mining companies and urban households. J Mod Afr Stud. 2018; 56(1): 113–41.

70. Serrazina B. Crossed cultures in Lunda, Angola: Diamang’s urban project and its legacies. Traditional Dwellings and Settlements Review. 2020; 31(2): 23–34.

71. Priebe GE, Macama A, Kessel B, dos Reis FVD, Van Dúnem JE, Katito J, et al. Associations between social capital and other living conditions among young Angolan women: a cross-sectional study in three provinces. medRxiv:25320530 [Preprint]. 2025 [cited 2025-01-20]. 10.1101/2025.01.14.25320530.

72. Instituto Nacional de Estatística (INE). Resultados definitivos recenseamento geral da população da habitação de Angola 2014, Huambo; Luanda; Lunda Sul, Angola. 2016. Available from: https://www.ine.gov.ao/publicacoes/detalhes/ODI3Ng%3D%3D.

73. Grais RF, Rose AM, Guthmann JP. Don’t spin the pen: two alternative methods for second-stage sampling in urban cluster surveys. Emerg Themes Epidemiol. 2017; 4: 8.

74. World Health Organization (WHO). Immunization coverage cluster survey: reference manual. Geneva: Switzerland. 2005. Available from: https://healthcluster.who.int/publications/m/item/who-vaccination-coverage-cluster-surveys-reference-manual.

75. Pelikan J, Ganahl K, Van den Broucke S, Sørensen K. Measuring health literacy in Europe: introducing the European Health Literacy Survey Questionnaire (HLS-EU-Q). In: Okan O, Bauer U, Levin-Zamir D, Pinheiro P, Sørensen K, editors. International handbook of health literacy: research, practice and policy across the life-span. Bristol, UK: Policy Press; 2019. pp. 115–138.

76. Stormacq C, Wosinski J, Boillat E, Van den Broucke S. Effects of health literacy interventions on health-related outcomes in socioeconomically disadvantaged adults living in the community: a systematic review. JBI Evid Synth. 2020; 18(7): 1389–469.

77. Chinn D, McCarthy C. All Aspects of Health Literacy Scale (AAHLS): developing a tool to measure functional, communicative and critical health literacy in primary healthcare settings. Patient Educ Couns. 2013; 90(2): 247–53.

78. McClintock HF, Alber JM, Schrauben S, Mazzola CM, Wiebe D. Constructing a measure of health literacy in Sub-Saharan African countries. Health Promot Int. 2020; 35(5): 907–15.

79. Dodson S, Good S, Osborne RH. Health literacy toolkit for low- and middle-income countries: a series of information sheets to empower communities and strengthen health systems. New Delhi: World Health Organization, Regional Office for South-East Asia, 2015. Available from: https://iris.who.int/bitstream/handle/10665/205244/B5148.pdf?sequence=1.

80. Upadhyay UD, Danza PY, Neilands TB, Gipson JD, Brindis CD, Hindin MJ, et al. Development and validation of the Sexual and Reproductive Empowerment Scale for Adolescents and Young Adults. J Adolesc Health. 2021; 68(1): 86–94.

81. Fernandes ET, Dias AC, Ferreira SL, Marques GC, Pereira CO. Adaptação cultural e confiabilidade da Reproductive Autonomy Scale para mulheres no Brasil. Acta Paul Enferm. 2019; 32(3): 298–304.

82. World Health Organization (WHO). WHO multi-country study on women’s health and domestic violence against women: initial results on prevalence, health outcomes and women’s responses. 2005. Available from: https://iris.who.int/handle/10665/43309.

83. Ondjango Feminista. Women’s economic resistance, a daily struggle. TUBA! Report. Fourth edition. 2021. Available from: https://static1.squarespace.com/static/57c54852f5e231e61738ab8a/t/6045f80355292b2bd739a7ee/1615198216886/TUBA-Ed4-2020-MASTER_ENG_compressed.pdf.

